# Test-Negative Designs with Multiple Testing Sources

**DOI:** 10.1101/2025.04.26.25326477

**Authors:** Mengxin Yu, Nicholas P. Jewell

## Abstract

Test-negative designs, a form of case-cohort studies, have been commonly used to assess infectious disease interventions. Early examples of the design included the evaluation of seasonal influenza vaccine in the field. Recently, they have also been widely used to evaluate the efficacy of COVID-19 vaccines in preventing symptomatic disease for different variants (Evans and Jewell, 2021). The design hinges on individuals being tested for the disease of interest; upon recruitment, such individuals are subjected to a definitive test for the presence of the disease of interest (test-positives), or not (test-negatives) along with the determination of whether the individual has been exposed to the intervention under study (e.g. vaccination). In most early TND studies, individuals were tested because they were suffering from symptoms consistent with the disease in question, and the TND was a tool to reduce confounding due to healthcare-seeking behavior. However, in many cases such as COVID-19, and Ebola, testing results were available at healthcare facilities for individuals who presented for a variety of reasons in addition to symptoms (e.g. case contact tracing, etc.). Aggregating samples from symptomatic and asymptomatic test results leads to bias in the assessment of the efficacy of the intervention. Here we consider these issues in the context of a specific version of the ‘multiple reasons for testing problem,’ motivated by a vaccine trial designed to assess a new Ebola viral disease vaccine (Watson-Jones et al., 2022). Some participants are recruited in the usual TND fashion as they present for care suffering from symptoms consistent with an Ebola diagnosis (and are thus tested); in addition, however, any test-positive identified in this fashion leads to immediate testing for Ebola for all close contacts of the test-positive who likely are asymptomatic at that point. We examine a simple approach to estimate a common efficacy of the vaccine intervention based on these two sources of test positives and test negatives, complemented with an assessment of whether efficacy is the same for both sources.

## 1. Introduction

The test-negative design (TND) has become common in studies of infectious disease interventions, and many TND studies have informed policy in the past decade. For example, policies informed by TND studies have included annual seasonal influenza vaccine recommendations (Grohskopf et al., 2019), implementation of a three-dose pneumococcal conjugate vaccine (PCV) schedule in the UK and other countries (Whitney et al., 2014), recommendations to remove upper age restrictions for receipt of rotavirus vaccines among children, and administration of oral cholera vaccines as a single dose during campaigns in emergency settings (icg, 2017; Hsiao et al., 2017; Qadri et al., 2018).

The TND is a version of a case-cohort study in which symptomatic patients meeting predefined inclusion/exclusion criteria are enrolled and subsequently classified as test-positive “cases” or test-negative “controls” based on the results of definitive diagnostic testing for the outcome under study. At the same time, eligible participants are classified as exposed or unexposed to an intervention of interest such as vaccination. As noted, this design is frequently used for evaluating the effectiveness of seasonal influenza vaccination (Jackson and Nelson, 2013; Foppa et al., 2013; Sullivan et al., 2016; Grohskopf et al., 2019), and its internal validity has been explored in depth (Orenstein et al., 2007; Jackson and Nelson, 2013; De Serres et al., 2013; Sullivan et al., 2014; Haber et al., 2015; Sullivan et al., 2016; Westreich and Hudgens, 2016). Briefly, validity depends primarily upon the avoidance of selection bias in the sampling of cases and controls, as well as the the extent to which the exposure distribution among controls is representative of the exposure distribution among the source population that gives rise to cases. Key assumptions and their relevance to test-negative studies are discussed briefly below. For ease of discussion we refer to the outcome of those testing positive for the pathogen of interest as “test-positive illness” and in those testing negative as “test-negative illness.”

The key assumptions for unbiased estimation of an intervention effect using data from a standard TND study include that (i) test-negative illness occurrence is not associated with the intervention, (ii) the relative propensity of individuals to seek health care is non-differential by outcome status, (iii) the test used to determine disease status is highly sensitive and specific, i.e. no misclassification, (iv) the sampling of test-negatives is unfiltered; that is, controls must be sampled from the whole population at risk without excluding those who test-positive at another time during the study period, (v) the efficacy of the intervention is not modified by healthcare-seeking behavior, and (vi) participants with test-negative illness are recruited only when test-positive illness is circulating. The first of these assumptions requires a careful definition of what is considered to be a suitable test-negative illness that also must exhibit non-specific symptoms similar to those that occur with the test-positive illness. Assumption (ii) is plausible if tested individuals are not aware of their (likely) outcome status prior to seeking care and testing. Assumption (iii) can be weakened by appropriate statistical adjustments. Assumptions (iv) and (vi) guarantee that test-negative controls are from the source population *at risk* for the test-positive illness. Finally, assumption (v) allows extrapolation from the healthcare-seeking population to the general population.

Because cases (test-positives) and controls (test-negatives) are recruited from the same patient population, and restricted to those seeking care, the design was intended to ameliorate if not eliminate, bias caused by health-care-seeking behavior (Jackson and Nelson, 2013; Haber et al., 2015; Sullivan et al., 2016).

Several authors have explored the statistical rationale and underlying assumptions of the TND, showing that the odds ratio (OR)—i.e., the odds of exposure in test-positives versus that for test-negative controls—is directly equivalent to the Relative Risk (RR) of the test-positive outcomes comparing exposed to unexposed *in the healthcare-seeking population*, providing underlying assumptions are met (Jackson and Nelson, 2013; Haber et al., 2015).

This is a direct consequence of the case-cohort nature of the sampling of test-positive and test-negative participants. We emphasize here that this Relative Risk is specific to the definition of a test-positive outcome in the sense that it depends on the level of symptoms that are required to be eligible for recruitment. As such, this Relative Risk may vary if eligibility requires more serious symptoms such as hospitalization, etc. In addition, the causal structure underlying this approach has recently been studied in detail using directed acyclic graphs and causal inference ideas (Sullivan et al., 2016). For the traditional TND examining influenza, exposure refers to seasonal vaccination so that an estimate of the RR for the outcome associated with exposure directly yields an estimate of vaccination efficacy, 1 − *RR*. For convenience, we shall denote the true comparative population RR by *λ*, again with the understanding that this efficacy is measured by a reduction in the likelihood of developing the specified symptoms necessary for recruitment.

Throughout the statistical development described above, it has been assumed that both test positives and test negatives are ascertained through healthcare facilities where potential participants seek care because of undifferentiated symptoms so that such individuals are blinded to their disease status at the time of seeking care when their exposure status (i.e. vaccination) is also determined. However, in recent applications, this simple ascertainment process is not satisfied in that there may be multiple reasons why individuals seek testing (that ultimately determines whether they are classified as test-positives or test-negatives). For example, in recent Covid-19 test-negative studies (Andrejko et al., 2022), individuals may seek, or be referred for, asymptomatic testing for screening purposes (for travel, etc), or because these individuals may have been exposed to the virus (by a family member, for example).

Lewnard et al. (2021) pointed out that reasons for testing other than symptoms may introduce bias, and Shi et al. (2023) conceptualized this issue through a causal graph. Both papers, and Vandenbroucke et al. (2022), state that a stratified analysis based on testing reasons could alleviate the bias, but no quantification or formal solution was given.

Here we focus on a specific example of a test-negative design with two distinct sources of participants, motivated by a proposed study to assess a vaccine for Ebola virus disease (EVD) in the eastern Democratic Republic of Congo (Watson-Jones et al., 2022; WHO, 2019; SAGE, 2019; Johnson and Johnson, 2019). In this trial, a traditional test-negative design was employed but, in addition to traditional symptomatic testing, close contacts of any known test-positives were all to be tested for the presence of the Ebola virus, prior to the onset of any symptoms. Vaccination status was ascertained for both symptomatic and asymptomatic participants at the time of recruitment.

Pearson et al. (2022) quantify the bias from a naive TND analysis based on data from this kind of hybrid design with two distinct reasons for testing: (i) symptomatic self-reporting or (ii) testing close contacts of known cases, and proposed a weighted average estimator to remove bias from naive aggregation. This required a strong assumption that the VE for either recruitment population is the same. Finally, a naive solution to filter out asymptomatic subjects and only use the remaining data to perform classical TND analysis was discussed. Although this procedure is valid, it loses a large amount of information, yielding less efficient estimation. In this paper, we discuss the analysis of data from such hybrid designs and examine a simple method for assessing the shared efficacy of vaccine intervention in response to these two sources (symptomatic samples and case contact samples) of test positives and test negatives, together with an assessment of whether the efficacy is the same for both sources. The merits of our method are shown by comparing results with existing benchmark estimators.

## 2. Traditional Test-Negative Data Supplemented by Testing of Close Contacts of Cases

Before discussing causal structures underlying the recruitment of both symptomatic and asymptomatic participants, we discuss briefly the role of an intervention such as vaccination in both preventing infections and specific symptoms. Following Lewnard et al. (2021), let *λ*_*S*_ be the relative risk of *infection* for a vaccinated versus unvaccinated individual, resulting from a reduced susceptibility to the acquisition of a pathogen or accelerated clearance of the pathogen. Further, define *λ*_*P*_ as the relative risk of the infection meeting a particular severity threshold, e.g., clinical symptoms requiring care, hospitalization, intensive care unit admission, or mechanical ventilation) for an infected vaccinated individual versus an infected unvaccinated individual (where *P* stands for progression). Then the overall relative risk (for the pre-defined symptom severity outcome) associated with vaccination is just *λ* = *λ*_*S*_ *× λ*_*P*_, with *V E* = 1 − *λ*. As we discuss below, the traditional TND (with symptomatic recruits) estimates *λ* but cannot separate out the distinct vaccine contributions *λ*_*S*_ and *λ*_*P*_. Further, this Relative Risk is estimated only in the healthcare-seeking population from which participants are drawn absent assumption (v) discussed in the previous section–for clarity, we refer to this Relative Risk as *λ*_*H*_. However, as we elucidate below, testing of case contacts provides direct estimates of *λ*_*S*_—which may differ from *λ*—in the entire population, whether healthcare-seekers or not. In fact, *λ* = *λ*_*S*_ if *λ*_*P*_ = 1, that is, the intervention— e.g. vaccination—has no influence on symptoms after infection has occurred. In short, for *λ*_*H*_ = *λ*_*S*_, we require—at least—assumption (v) above and that *λ*_*P*_ = 1. We discuss additional confounding below.

We now elucidate this reasoning by exploiting two Directed Acyclic Graphs (DAGs) associated with test-negative designs for the two distinct reasons for testing.

### 2.1 Test-Negative Design with Symptomatic Participants

The illustration on the left panel of Figure 1 describes a DAG for a traditional test-negative design that recruits symptomatic patients only. These are individuals who proactively seek medical attention when they experience symptoms pertinent to the disease of concern (and the test-negative conditions for that matter). Within this population, the results of an appropriate diagnostic test are classified into two groups: ‘test-positive’ and ‘test-negative’ (control).

**Figure 1.**
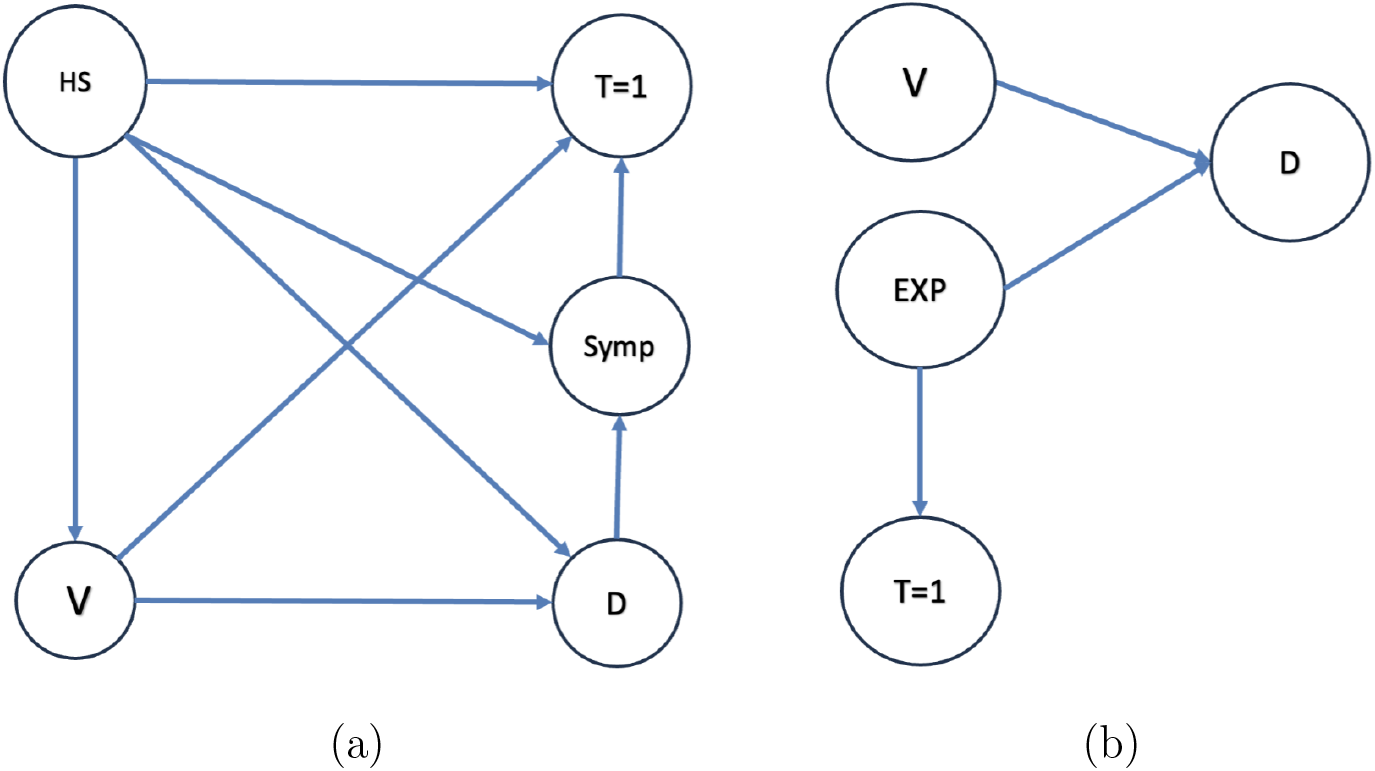
DAGs for Self-Reported (Symptomatic) and Case Contact Tracing (Asymptomatic)

**Figure 2.**
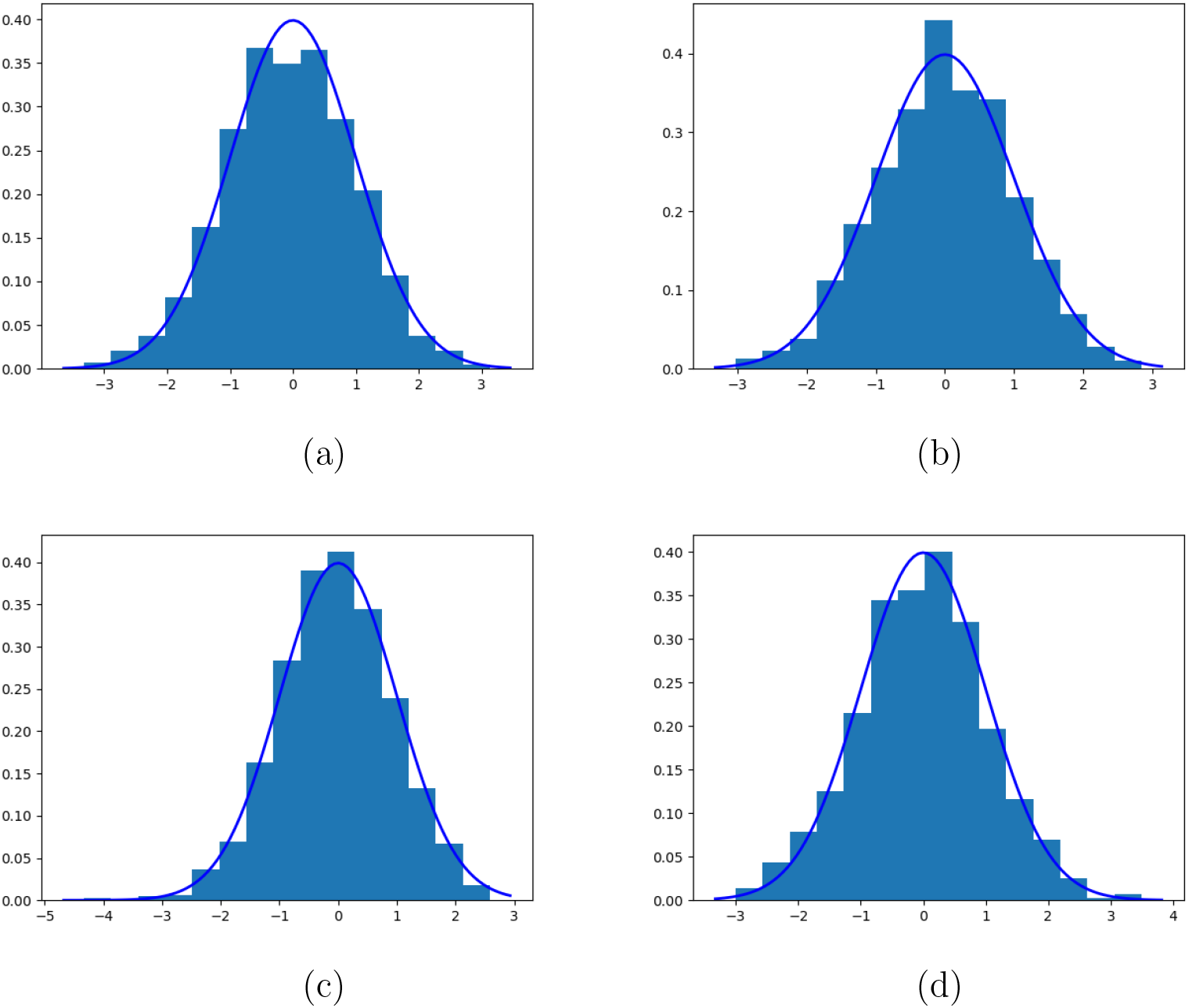
Asymptotic distribution of normalized 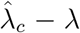 under different cases. (a): *λ* = 1.0, *n* = 500; (b): *λ* = 1.0, *n* = 1000; (c): *λ* = 0.9, *n* = 500; (b): *λ* = 0.9, *n* = 1000;

**Figure 3.**
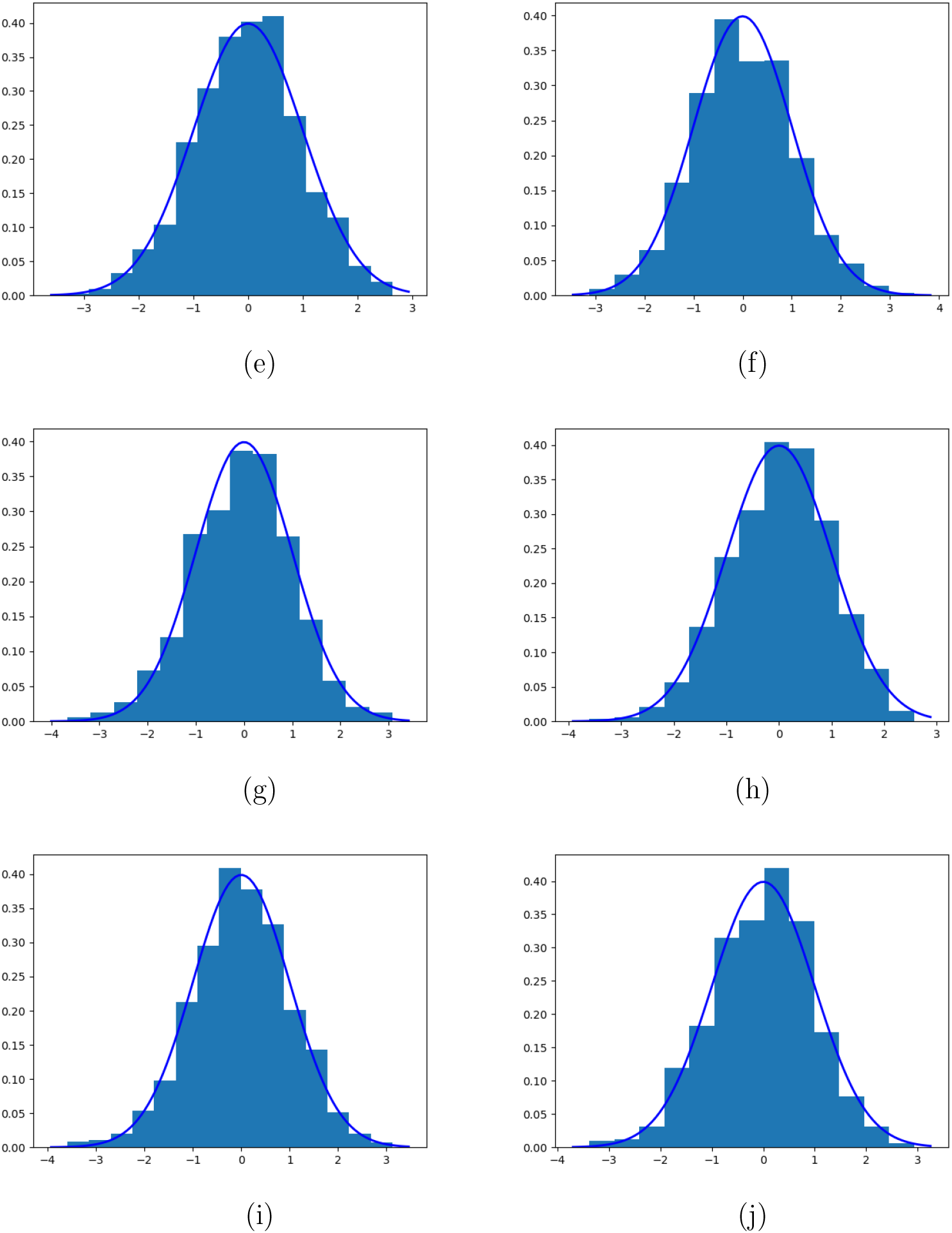
Asymptotic distribution of normalized 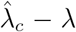 under different cases. (e): *λ* = 0.8, *n* = 500; (f): *λ* = 0.8, *n* = 1000; (g): *λ* = 0.7, *n* = 500; (h): *λ* = 0.7, *n* = 1000; (i): *λ* = 0.6, *n* = 500; (j): *λ* = 0.6, *n* = 1000;

We define the several nodes, or variables, within this DAG, as follows:

- *HS*: This node denotes the variable associated with quantifying Healthcare-Seeking behavior. For simplicity, we treat this variable as binary. Specifically, *HS* = 1 indicates that the participant seeks care upon onset of the symptoms defined by the study protocol. Only such individuals can be recruited in a traditional test-negative study.
- V: This node refers to the vaccination, or intervention, status of the patient.
- D: This node signifies the disease status, elucidating whether the target infection has occurred or not (i.e. a test-positive result).
- Symp: This node is an indicator variable for the presence of disease-related (test-positive or test-negative) symptoms.
- *T* = 1: This depicts whether a test has been conducted, essentially that the participant is eligible and has been recruited to the study.

The arrow *V* → *D* is indicative of the fact that a patient’s vaccination status can influence the likelihood of contracting the disease in question. There is no direct effect from *D* → *T* since taking the test is entirely determined by health-seeking behavior and symptoms.

Next, we introduce an additional assumption regarding the relationship between vaccine effectiveness, healthcare-seeking behavior, and other (test-negative) illnesses that permit identification.

Assumption 1: We posit that the effectiveness of the vaccine is not modified by healthcare-seeking behavior and the vaccine status is also independent of the presence, or not of the test-negative illnesses.

The second part of this assumption (discussed as Assumption (i) in Section 1) permits the identification of vaccine effectiveness, whereas the first part allows extrapolation of estimated vaccine effectiveness to the entire symptomatic population whether they seek care or not. These assumptions are also made in many previous test-negative design papers (Jackson and Nelson, 2013; Pearson et al., 2022). Note that the traditional test-negative design cannot assess vaccine effectiveness in preventing infection, only on preventing *symptomatic* infection. We will discuss this later when we expand the design to include testing asymptomatic participants.

Identification of vaccine effectiveness within the symptomatic tested population then follows:

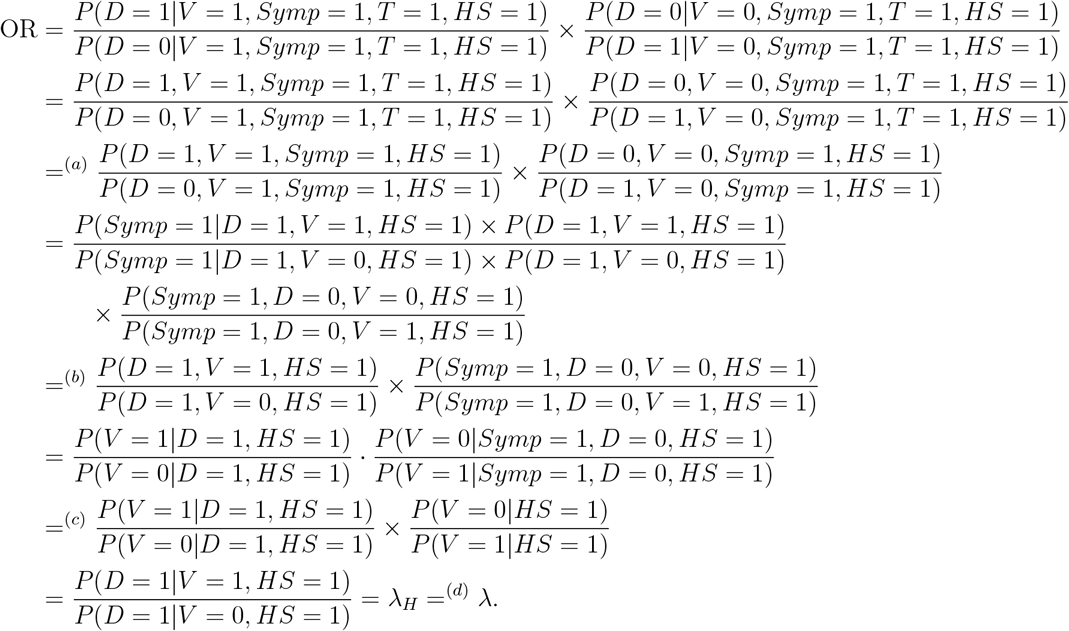

There are several steps listed above that merit further explanation. Specifically, equality (*a*) holds since

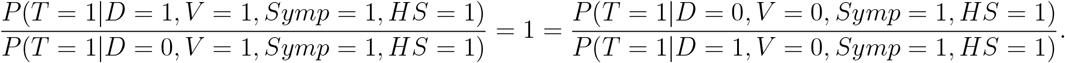

Essentially, with a given vaccine status *V Symp* = 1, and healthcare-seeking behavior *HS* = 1, the decision to test *T* remains independent of the disease status *D*. This is a valid observation in the context of the symptomatic healthcare-seeking population, where the decision to pursue testing is primarily influenced by healthcare-seeking behavior, symptoms, and vaccine status. Disease status only influences testing behavior indirectly, through the manifestation of relevant symptoms. Note that in well-designed test-negative studies, this assumption is reasonable since participants do not know their exact disease status before testing. The assumption might be violated if, for example, other circumstances suggest disease status to the participant before testing (such as knowledge of close contact with the relevant pathogen of interest).

Equality (*b*) holds since

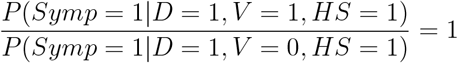

The underlying logic of this equation is that given a certain health-seeking behavior *HS* and disease outcome *D*, the status of symptoms is directly dictated by the presence of disease (*D* = 1), and remains unaffected by vaccine status. This directly assumes that the vaccine does not prevent (or ameliorate) symptoms, or in symbols that *λ*_*P*_ = 1; however, if infected by the test-positive pathogen, it is possible that prior vaccination may reduce or even eliminate symptoms (as has been postulated for Covid-19 vaccines). This is unlikely to be true for vaccines for Ebola viral disease, however.

Equality (*c*) is valid since per Assumption 1, amongst control participants (i.e. test-negatives), the vaccine status is independent of symptoms, and its relative frequency reflects that of the general healthcare-seeking population.

Finally, Assumption 1 posits that the result of the intervention is not modified by the health-seeking behavior, validating Equality (*d*). Consequently, we derive the following expression:

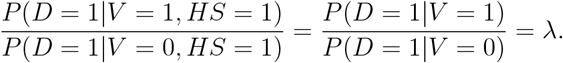

Based on the identification assumptions, we are thus able to leverage the symptomatic participant data on vaccination to ascertain the true effectiveness of the vaccine. We now briefly outline the sampling context for the traditional test-negative design used for recruiting individuals. Table 1 provides the breakdown of both test-positives (*D*) and test-negatives (*D*^*c*^) by their vaccination status. This yields the following (conditional) likelihood, where we use the subscript *SR* to denote the data arising from symptomatic recruited patients:

**Table 1.** Test samples from the self-reported population. In this context, the rows D and D^c^ represent test-positive and test-negative groups, respectively. The columns V and V ^c^ denote whether individuals within a specific group (positive or negative) have been vaccinated.

|  |  |  |
| --- | --- | --- |
| | $V$ | $V^c$ |
| $D$ | $A'$ | $C'$ |
| $D^c$ | $B'$ | $D'$ |

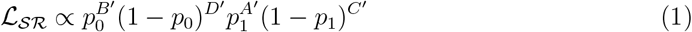

where *p*_0_ = *p*(*V*) and *p* = *P* (*V D*) with 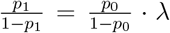. In this context, *A*′, *B*′, *C*′, and *D*′ represent the number of observed test samples in distinct categories from symptomatic participants.

### 2.2 Testing of Contacts of Positives (Cases)

The right-hand panel of Figure 1 illustrates a Directed Acyclic Graph (DAG) for data that arises from testing close contacts of test-positives. In this population, the testing behavior, as per the DAG, is solely determined by the exposure status (*EXP*), where *EXP* = 1 indicates that an individual is a close contact with a known person who has tested positive for the presence of *D*; the decision to test is unaffected by the individual’s vaccine status (*V*) and disease status (*D*), given *EXP* = 1. Such a DAG might describe scenarios involving severe diseases, such as Ebola, where individuals who have had close contact with confirmed positive cases are expected to undergo immediate (asymptomatic) testing. Consequently, we make the following assumption:

Assumption 2: Any individual identified as a close contact is obligated to undertake a screening test for the presence of *D*, that is,

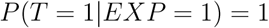

This assumption follows the protocol of the Ebola vaccine trial (Pearson et al., 2022).

Next, we propose an additional assumption that the efficacy of the vaccine is independent of the exposed population.

Assumption 3: The efficacy of the vaccine is independent of exposed status.

Under these two assumptions, using *RR* to denote the relative frequency of positive test results among the tested contacts, we have

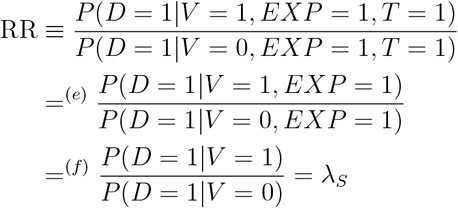

Equality (*e*) follows from Assumption 2, which posits that all close contacts undergo testing given exposure to confirmed positive cases. Furthermore, Equality (*f*) is valid due to Assumption 3, which assumes that the efficacy of the vaccine is independent of exposure status. Note that, in this context, the variable *D* = 1 requires only a positive test for infection with the pathogen and does not require the presence of any symptoms. Recall that *λ*_*S*_ = *λ* if *λ*_*P*_ = 1.

Given this identifiability, we consider the following (conditional) likelihood arising from Table 2 data that classifies contacts’ test results by outcome and vaccination status of the contact:

**Table 2.** Test sample counts from testing close contacts of positives. The definition of D, D^c^, V, V ^c^ are consistent with those in Table 1

| | $V$ | $V^c$ |
| --- | --- | --- |
| $D$ | $A^*$ | $C^*$ |
| $D^c$ | $B^*$ | $D^*$ |

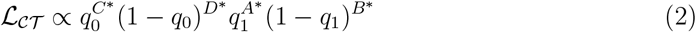

where 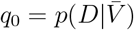and *q*_1_ = *P* (*D*|*V*) = *λ*_*S*_*q*_0_.

### 2.3 Combined Likelihood

From the discussion of Sections 2.1 and 2.2, we note the presence of two distinct likelihood functions, each capable of evaluating the effectiveness of the vaccine using observed samples, albeit estimating somewhat different measures of efficacy absent further assumptions. If we invoke the assumptions discussed above, including that *λ*_*P*_ = 1, and we assume that the two estimands are equivalent for this and other reasons (we turn to a discussion of confounding later in this section), we propose a straightforward methodology to assess the collective efficacy of the vaccine intervention by aggregating the two distinct sources of information. If we use a composite likelihood, the full (conditional likelihood is then simply ℒ = ℒ _*𝒮 ℛ*_ *×*ℒ _*𝒞 𝒯*_.

When contacts of cases from the initial TND are tested (as was planned for the motivating Ebola trial), the use of this combined likelihood makes an additional assumption, namely that the outcome of a case contact is independent of the characteristics of the associated case;

The negative logarithm of this combined likelihood ℒ = ℒ _*SR*_ *×*ℒ _*CT*_ simplifies to

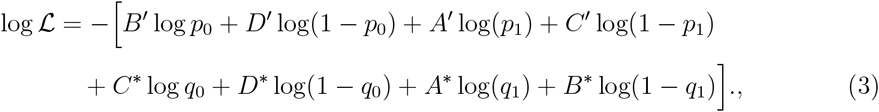

based on the data from Tables 1 and 2. Furthermore, from sections 2.1 and 2.2, we substitute *p*_1_ with *p*_0_*λ/*(1 − *p*_0_ + *p*_0_*λ*) and replace *q*_1_ with *λq*_0_. Our primary interest lies in conducting statistical inference for the common *λ*.

We note that the parameters *p*_0_, *p*_1_, *q*_0_, *q*_1_ are constrained within the interval [0, 1], whereas the parameter *λ* is confined to the non-negative real numbers. These restrictions pose challenges for optimization procedures, due to the necessity of operating within a constrained parameter space. To address this, we use a simple reparameterization. Specifically, we redefine *p*_0_ as 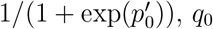 as 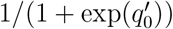,and *λ* as exp(*λ*′).

Then, employing this reparameterization in the combined likelihood (3) yields

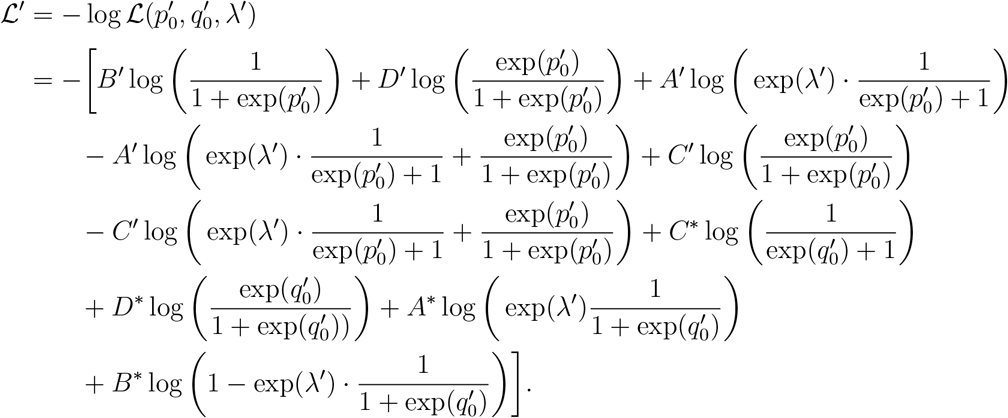

Subsequently, assuming that the true value of (*p*_0_, *q*_0_, *λ*) does not lie on the boundary of the parameter space, it is then straightforward to establish the existence of a unique maximum likelihood estimate, denoted by 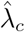.

We end this section by noting that, of course, the two likelihood functions (1) and (2) can be maximized separately to provide estimates of *λ* and *λ*_*S*_. With this approach, a likelihood ratio test can be used to test the equality of the two underlying Relative Risks. Note that rejection of the null hypothesis, in this case, does not formally establish that *λ*_*P*_ = 1 since there are other potential explanations of why the two Relative Risks may differ. For example, the exposure experiences of close contacts of cases may differ from those recruited symptomatically from the general population which may affect the performance of a vaccine intervention. Further, there is the issue of confounding by healthcare-seeking behavior amongst case contacts. Nevertheless, this likelihood ratio test may provide some evidence that a vaccine influences the progression of symptoms after breakthrough infections. To address potential confounding, it is important to expand the likelihood functions (1) and (2) to incorporate the influence of relevant covariates. The choice of covariates might differ for the distinct likelihoods as varying covariate information might be available from the two data sources. Particularly important here is the role of healthcare-seeking behavior.

This variable is naturally controlled in the estimation of *λ* from the first likelihood, ℒ = ℒ _*𝒮 ℛ*_. but is not addressed by naive use of ℒ = ℒ _*𝒮 ℛ*_ .In this case, measurement and adjustment for covariates that act as proxies for healthcare-seeking behavior may be especially important.

When covariates are included (through the use of appropriate GLMs in both likelihood components), the estimand reflects a conditional Relative Risk and thus vaccine effectiveness. Given the collapsibility of the Relative Risk, this will still yield a marginal Relative Risk associated with *V* so long as there is no interaction between *V* and any of the included covariates. Note that—for most vaccines—there is usually no biological basis for expecting such interactions to occur (other than perhaps for measures of immune compromise).

Of further interest is the vaccination status of the case associated with contacts in data underlying likelihood function (2). This information should be available for contacts of known cases (particularly if the cases arise from symptomatic recruitments in the underlying TND study). It is unlikely, however, that this is known for the TND participants since in most cases the source of their infection is likely unknown. Analysis of this covariate in an expanded version of (2) would allow assessment of whether vaccination reduces infectiousness in a vaccinated individual who experiences a breakthrough infection.

Note that adjustment for covariates may also control for any source of dependence between a case (used in likelihood (1)) and contacts (used in likelihood (2)) which has been ignored in use of the combined likelihood as noted above.

## 3. Numerical Studies

In this section, we assess the performance of the combined maximum likelihood estimator through simulations. For simplicity, we consider sample sizes of *n* = 500 with (*B*′ + *D*′) = (*A*′ + *C*′) = (*C*^*^ + *D*^*^) = (*A*^*^ + *B*^*^) = *n*.

We selected the true parameters 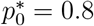 and 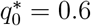 while varying the value of *λ* across the range 0.6 ∼ 1.0, reflecting high rates of vaccination and moderate values of efficacy. We provide a detailed implementation of the optimization procedure in the appendix.

We compare the performance of the estimator 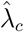,obtained by minimizing the combined loss function in (3), with additional benchmark estimators. In simulating the data, we allow for two situations: (i) the vaccine effects of the two sources are the same and (ii) vaccine effects from the two sources are different, for example, whether *λ* = *λ*_*S*_ or not.

For each case, we study four benchmarks involving different weighted averages of the two estimators based on separate maximizations of (1) and (2), denoted as 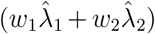,where 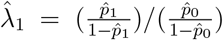 and 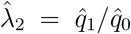.Here, 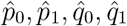 represent the separate maximum likelihood estimators based on (1) and (2), respectively. It is noteworthy that a weighted estimator was previously suggested by Pearson et al. (2022); however, the specific choice of weights was not explicitly discussed. Therefore, we compare estimation results using 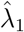 with (i) the traditional TND estimator where we set *w*_1_ = 1 (benchmark 1) and (ii) a simple average (with *w*_1_ = 1*/*2, *w*_2_ = 1*/*2) (benchmark 2) and (iii) a third benchmark (benchmark 3) where the weights are proportional to the (estimated) inverse standard deviations of 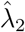 and 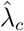.In addition, we also compare 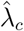 with an odds ratios estimator based on a naive aggregation of the two distinct testing samples from the two sources (the fourth benchmark); that is, the estimator [(*A*′ + *A*^*^) *×* (*D*′ + *D*^*^)]*/*[(*B*′ + *B*^*^) *×* (*C*′ + *C*^*^)].

For each assumed value of *λ*, we generate 500 simulated data sets, reporting the average value of our estimator 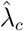 as well as the standard error of 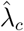.Detailed simulation results are presented in Table 3. We also consider scenarios where the vaccine effects from the two testing sources are different (e.g. when *λ* ≠ *λ*_*S*_). In this case, we study the performance of the aforementioned estimators and also conduct a likelihood ratio test with significance level *α* = 0.05 on the equivalence of the vaccine effect from two sources. The results are presented in Tables 4 and 5, respectively.

**Table 3.** Results on the estimation of vaccine effectiveness (1 − λ) when the two testing sources share the same vaccine effect. Proposed Estimator: The estimator maximizes the combined likelihood. Benchmark 1: the traditional TND estimator based on symptomatic participants only. Benchmark 2: Simple average of 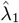 and 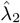.Benchmark 3: Weighted average of 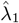 and 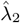,with weights proportional to the (estimated) inverse standard deviation of 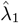 and 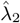,respectively. Benchmark 4: The odds ratio estimator aggregating data from the two testing sources (symptomatic and case contacts). SDs provide the observed standard deviations of the estimators over the 500 simulated samples

|  |  | Proposed Estimator | Benchmark 1 | Benchmark 2 | Benchmark 3 | Benchmark 4 |
| --- | --- | --- | --- | --- | --- | --- |
| $\lambda = 1.0$ | Averages | 1.001 | 1.019 | 0.879 | 1.010 | 1.005 |
|  | SDs | 0.049 | 0.163 | 0.085 | 0.055 | 0.069 |
| $\lambda = 0.9$ | Averages | 0.902 | 0.904 | 0.904 | 0.903 | 0.773 |
|  | SDs | 0.048 | 0.136 | 0.074 | 0.053 | 0.073 |
| $\lambda = 0.8$ | Averages | 0.799 | 0.808 | 0.804 | 0.802 | 0.673 |
|  | SDs | 0.043 | 0.122 | 0.065 | 0.047 | 0.060 |
| $\lambda = 0.7$ | Averages | 0.700 | 0.704 | 0.703 | 0.702 | 0.584 |
|  | SDs | 0.041 | 0.108 | 0.058 | 0.045 | 0.054 |
| $\lambda = 0.6$ | Averages | 0.601 | 0.609 | 0.605 | 0.603 | 0.500 |
|  | SDs | 0.038 | 0.092 | 0.051 | 0.041 | 0.047 |

**Table 4.** Results on the estimation of vaccine effectiveness (1− λ) when the two testing sources do not share the same vaccine effect. The vaccine effect in the TND population (symptomatic recruitment) takes values λ_1_≡ λ varying between 0.5∼ 0.9. In addition, we let λ_2_≡ λ_S_ = 1.0 be the Relative Risk amongst the case contact population. Proposed Estimator: The estimator maximizes the combined likelihood. Benchmark 1: the traditional TND estimator based on symptomatic patients only. Benchmark 2: Simple average of 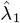 and 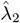.Benchmark 3: Weighted average of 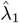 and 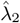,with weights proportional to the (estimated) inverse standard deviation of 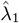 and 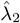 respectively. Benchmark 4: The odds ratio estimator aggregating data from the two testing sources (symptomatic and case contacts). SDs provide the observed standard deviations of the estimators over the 500 simulated samples

|  |  | Proposed Estimator | Benchmark 1 | Benchmark 2 | Benchmark 3 | Benchmark 4 |
| --- | --- | --- | --- | --- | --- | --- |
| $\lambda_1 = 0.9$ | Averages | 0.979 | 0.910 | 0.956 | 0.977 | 0.849 |
|  | SDs | 0.049 | 0.145 | 0.078 | 0.055 | 0.079 |
| $\lambda_1 = 0.8$ | Averages | 0.963 | 0.801 | 0.900 | 0.941 | 0.811 |
|  | SDs | 0.047 | 0.122 | 0.067 | 0.051 | 0.075 |
| $\lambda_1 = 0.7$ | Averages | 0.959 | 0.702 | 0.851 | 0.901 | 0.775 |
|  | SDs | 0.045 | 0.101 | 0.056 | 0.047 | 0.068 |
| $\lambda_1 = 0.6$ | Averages | 0.943 | 0.605 | 0.804 | 0.856 | 0.735 |
|  | SDs | 0.047 | 0.090 | 0.052 | 0.048 | 0.069 |
| $\lambda_1 = 0.5$ | Averages | 0.917 | 0.504 | 0.751 | 0.795 | 0.679 |
|  | SDs | 0.046 | 0.074 | 0.047 | 0.046 | 0.063 |

**Table 5.** Results of Likelihood Ratio Test for λ_1_ ≡λ = λ_2_ ≡λ_S_ at significance level α = 0.05. The Relative Risk of the case contact population is set at λ_2_ ≡λ_S_ = 1.0, with the Relative Risk of the TND population (symptomatic recruitment) λ varying among {0.4, 0.6, 0.8, 1.0}. Here, overall sample size is determined by n = (B′ + D′) = (A′ + C′) = (C^*^ + D^*^) = (A^*^ + B^*^).

|  |  | Power or Type-I Error |
| --- | --- | --- |
| $n = 200$ | $\lambda_2 = 1.0, \lambda_1 = 1.0$ | 0.049 |
| | $\lambda_2 = 1.0, \lambda_1 = 0.8$ | 0.164 |
| | $\lambda_2 = 1.0, \lambda_1 = 0.6$ | 0.636 |
| | $\lambda_2 = 1.0, \lambda_1 = 0.4$ | 0.968 |
| $n = 400$ | $\lambda_2 = 1.0, \lambda_1 = 1.0$ | 0.049 |
| | $\lambda_2 = 1.0, \lambda_1 = 0.8$ | 0.268 |
| | $\lambda_2 = 1.0, \lambda_1 = 0.6$ | 0.827 |
| | $\lambda_2 = 1.0, \lambda_1 = 0.4$ | 1.000 |
| $n = 600$ | $\lambda_2 = 1.0, \lambda_1 = 1.0$ | 0.045 |
| | $\lambda_2 = 1.0, \lambda_1 = 0.8$ | 0.341 |
| | $\lambda_2 = 1.0, \lambda_1 = 0.6$ | 0.943 |
| | $\lambda_2 = 1.0, \lambda_1 = 0.4$ | 1.000 |

### 3.1 Results

Based on Table 3, the simple aggregated odds ratio (without differentiating the testing sources/reasons; that is, Benchmark 4) exhibits bias. This occurs even though the vaccine effects from the two distinct testing sources are identical, leading to a failure to accurately identify the true vaccine effect. In contrast, when data from the two testing sources are separately analyzed, our proposed estimator—derived from the combined likelihood of Equation 3—demonstrates greater efficiency as compared to the other benchmark estimators. This assertion is further corroborated by the theoretical findings presented in the appendix, wherein we establish that the variance of the proposed estimator achieves the Cramér–Rao lower bound.

Table 4 reveals that when the vaccine effects from the two testing sources differ (*λ* ≠ *λ*_*S*_), all combined estimators exhibit bias. Note that the simulations here reflect a possible scenario where *λ*_1_ *< λ*_2_ = 1, so the vaccine here does not reduce the risk of infection but reduces the risk of symptoms post-infection. Therefore, in reality, it is important to first assess the comparability of vaccine effects from the distinct data sources, for example by implementing a likelihood ratio test. To this end, we present the results of conducting such a likelihood test in Table 5. When the null hypothesis holds, the type I error is well controlled under 5%, and when the alternative holds, the power rapidly increases to 1 when the number of testing samples grows.

## 4. Discussion

Our analysis of the Ebola vaccine study example with two sources of testing data illustrates the importance of considering the reasons for testing and avoiding any temptation to aggregate testing data naively. Further, it is crucial to understand that data arising from different testing sources may estimate different measures of effectiveness. The design of such hybrid TND studies must carefully consider an appropriate discussion of symptoms for eligibility as well as the possibility that the intervention may alleviate the development of such symptoms separately from any effect on infection. In most cases, it will also be important to pre-define covariates for measurement that will allow adjustment for potential confounding by healthcare-seeking behavior when implementing asymptomatic testing. When the estimands associated with different testing sources differ, further information will usually be required to disentangle the root causes of such differences.

## Data Availability

All data produced in the present study are available upon reasonable request to the authors

## 5. Acknowledgment

This research was supported by grant #R01-AI148127 from the National Institute for Allergy and Infectious Diseases. The authors would like to thank Dylan S. Small and Bingkai Wang for their insightful comments and discussion on this paper.

## Algorithm

Notably, if we independently solve the two distinct loss functions presented in sections 2.1 and 2.2, respectively, we arrive at a consistent estimator for the parameters 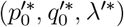.This consistent estimator can be selected as the initialization point for our gradient descent algorithm. This strategy leads to the first phase of the algorithm, which is concerned with initialization, as detailed in the subsequent discussion.:

Step 1, Initialization: We first solve 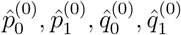 by maximizing likelihoods (1) and (2), respectively. We then initialize the re-parameterized parameters at the zeroth (initial) step 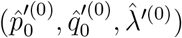.In specific, we first let 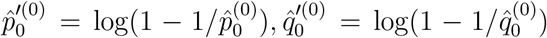 and initialize the estimator of 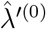 by an log- weighted average of 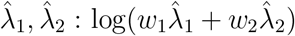, where 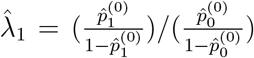 and 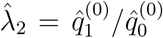.The weights *w*_1_, *w*_2_ with *w*_1_ + *w*_2_ = 1 are pre-determined. It can be chosen equally or proportional to the standard deviation of 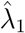 and 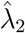,respectively.

Following the initialization phase, we progress to the subsequent stage, which entails the execution of the gradient descent algorithm. The specifics of this step are elucidated in the following description.

Step 2, Run Gradient Descent: Choose stepsize *η* and run gradient descent on the parameters 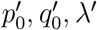. The updates are presented as follows:

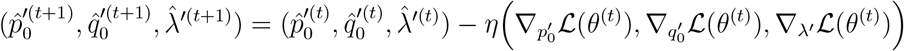

where 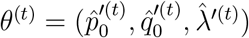.The gradient is given as follows:

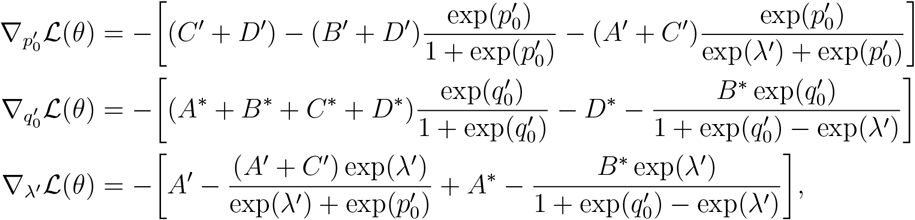

where 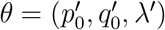.

Step 3, Stopping Criterion: Upon reaching a point in the iterative process denoted as *θ*^(*t*)^, we terminate the algorithm when the infinity norm of the gradient at that particular iterate falls below a predetermined threshold value, denoted as *τ*.

In the simulation experiments, we choose a step size of *η* = 0.01*/n*, and a stopping threshold of *τ* = 10^−5^ is used.

## Theoretical Guarantees

In the previous simulation example, we observed that our estimator outperformed weighted alternatives in terms of both finite sample bias and standard deviation.

By investigating the theoretical guarantee of the asymptotic variance, we aim to provide a deeper understanding of the statistical properties and performance limits of our proposed estimator. This analysis will shed light on the precision and reliability of our estimator as sample sizes grow and allow for valuable insights into its practical applications.

### Uncertainty Quantification and Cramer Rao Lower bound

Since our joint estimator 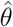minimizes the negative log-likelihood (3), according to Taylor Expansion, we know

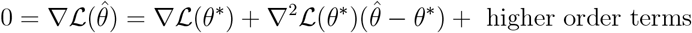

where 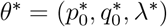.By direct calculation, we obtain

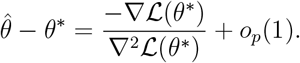

All involved distributions are binary distributions with bounded support. Moreover, the density function is also continuous to *θ* = (*p*_0_, *q*_0_, *λ*). Therefore, the regularity condition holds and Var(*∇ℓ*(*θ*^*^)) = 𝔼 [*∇*^*2*^*ℓ*(*θ*^*^)]. (since we are studying the negative log-likelihood)

Theorem 1: *We have* 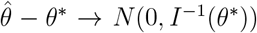 *where I*(*θ*^*^) = 𝔼 [*∇*^*2*^*ℓ*(*θ*^*^)] *is the Fisher information matrix evaluated under θ*^*^.

Based on the presented theorem, we can deduce that our joint estimator has the capability to achieve the minimum variance among all consistent estimators. This finding provides a theoretical validation for our previous observation that our estimator exhibits superior efficiency in terms of standard deviation. It confirms that our estimator possesses favorable statistical properties, reinforcing its potential as an optimal choice for parameter estimation in the given context.

Next, we use some simulations to validate the asymptotic variance in the following section.

## Simulation Validation

In this section, we validate the asymptotic variance of 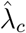 presented in Theorem 1. n this simulation, we employ the same settings as outlined in Section 3.1. We repeat the experiments 1000 times and record the normalized estimators, which are obtained by dividing the estimators by the corresponding variances presented in Theorem 1, for each replication. The resulting normalized estimators are then used to construct histograms, which are presented below.

The presented figures consist of two panels, corresponding to experiments conducted with sample sizes of *n* = 500 and *n* = 1000, respectively. Each figure contains five rows, representing the results obtained for different true parameter values *λ* = 1, 0.9, 0.8, 0.7, 0.6.

These histograms provide an empirical visualization of the distribution of the normalized estimators, offering insights into the accuracy and precision of our proposed methodology under different true parameter settings and sample sizes.

## References

(2017). Provisional ICG for Oral Cholera Vaccine. Report of the Annual Meeting, Geneva. http://apps.who.int/iris/bitstream/handle/10665/259829/WHO-WHE-IHM-2017.13-eng.pdf?sequence=1.

Andrejko, K. L., Pry, J., Myers, J. F., Jewell, N. P., Openshaw, J., Watt, J., Jain, S., Lewnard, J. A., and the California COVID-19 Case-Control Study Team (2022). Prevention of coronavirus disease 2019 (covid-19) by mrna-based vaccines within the general population of California. Clinical Infectious Diseases, 74(8):1382–1389.

De Serres, G., Skowronski, D., Wu, X., and et al. (2013). The test-negative design: validity, accuracy and precision of vaccine efficacy estimates compared to the gold standard of randomised placebo-controlled clinical trials. Eurosurveillance, 18:3104–3109.

Evans, S. and Jewell, N. P. (2021). Vaccine effectiveness studies in the field. New England Journal of Medicine, 385:650–651.

Foppa, I., Haber, M., Ferdinand, M., and et al. (2013). The case test-negative design for studies of the effectiveness of influenza vaccine. Vaccine, 31:3104–3109.

Grohskopf, L., Alyanak, E., Broder, K., and et al. (2019). Prevention and control of seasonal influenza with vaccines: recommendations of the advisory committee on immunization practices—–United States, 2019-20 influenza season. MMWR Recomm Rep, 68:1–21.

Haber, M., An, Q., Foppa, I., and et al. (2015). A probability model for evaluating the bias and precision of influenza vaccine effectiveness estimates from case-control studies. Epidemiology & Infection, 143:1417–1426.

Hsiao, A., Desai, S., Mogasale, V., Excler, J., and Digilio, L. (2017). Lessons learned from 12 oral cholera vaccine campaigns in resource-poor settings. Bull World Health Organ, 95:303–312.

Jackson, M. L. and Nelson, J. C. (2013). The test-negative design for estimating influenza vaccine effectiveness. Vaccine, 31(17):2165–2168.

Johnson and Johnson (2019). Johnson & Johnson Announces Donation of up to 500,000 Regimens of Janssen’s Investigational Ebola Vaccine to Support Outbreak Response in Democratic Republic of the Congo (DRC). https://www.jnj.com/johnson-johnson-announces-donation-of-up-to-500-000-regimens-of-jansens-investigational-ebola-vaccine-to-support-outbreak-response-in-democratic-republic-of-the-congo-drc.

Lewnard, J. A., Patel, M. M., Jewell, N. P., Verani, J. R., Kobayashi, M., Tenforde, M. W., Dean, N. E., Cowling, B. J., and Lopman, B. A. (2021). Theoretical framework for retrospective studies of the effectiveness of sars-cov-2 vaccines. Epidemiology, 32(4):508–517.

Orenstein, E., De Serres, G., Haber, M., and et al. (2007). Methodologic issues regarding the use of three observational study designs to assess influenza vaccine effectiveness. International Journal of Epidemiology, 36:623–631.

Pearson, C. A., Edmunds, W. J., Hladish, T. J., and Eggo, R. M. (2022). Potential test-negative design study bias in outbreak settings: application to ebola vaccination in democratic republic of congo. International Journal of Epidemiology, 51(1):265–278.

Qadri, F., Azad, A., Flora, M., and et al. (2018). Emergency deployment of oral cholera vaccine for the rohingya in bangladesh. Lancet, 391:1877–1879.

SAGE (2019). Strategic Advisory Group of Experts (SAGE) on Im-munization Interim Recommendations on Vaccination against Ebola Virus Disease (EVD). https://www.who.int/immunization/policy/position_papers/interim_ebola_recommendations_may_2019.pdf.

Shi, X., Li, K. Q., and Mukherjee, B. (2023). Current challenges with the use of test-negative designs for modeling covid-19 vaccination and outcomes. American Journal of Epidemiology, 192(3):328–333.

Sullivan, S., Feng, S., and Cowling, B. (2014). Potential of the test-negative design for measuring influenza vaccine effectiveness: a systematic review. Expert Review of Vaccines, 13:1571–1591.

Sullivan, S., Tchetgen Tchetgen, E., and Cowling, B. (2016). Theoretical basis of the test-negative study design for assessment of influenza vaccine effectiveness. American Journal of Epidemiology, 184(5):345–353.

Vandenbroucke, J., Brickley, E., Pearce, N., and Vandenbroucke-Grauls, C. (2022). The evolving usefulness of the test-negative design in studying risk factors for covid-19. Epidemiology, 33(2):e7–e8.

Watson-Jones, D., Kavunga-Membo, H., Grais, R. F., Ahuka, S., Roberts, N., Edmunds, W. J., Choi, E. M., Roberts, C. H., Edwards, T., Camacho, A., et al. (2022). Protocol for a phase 3 trial to evaluate the effectiveness and safety of a heterologous, two-dose vaccine for ebola virus disease in the democratic republic of the congo. BMJ Open, 12(3):e055596.

Westreich, D. and Hudgens, M. G. (2016). Invited commentary: beware the test-negative design. American Journal of Epidemiology, 184(5):354–356.

Whitney, C., Goldblatt, D., and O’Brien, K. (2014). Dosing schedules for pneumococcal conjugate vaccine: considerations for policy makers. Pediatr Infect Dis J, 33(Suppl 2):S172–181.

WHO (2019). Weekly Epidemiological Record 2019. https://www.who.int/immunization/sage/meetings/2019/april/SAGE_April_2019_meeting_summary.pdf.

